# Factors Associated with the use of Long-Acting Reversible Contraceptives among adolescents at first level hospitals in Lusaka, Zambia

**DOI:** 10.64898/2026.08.26.26361478

**Authors:** Imanga Ikabongo, Swebby Macha, Bellington Vwalika, Patrick Kaonga, Maureen M. Masumo, Hikabasa Halwiindi, Evaristo Kunka, Alice Ngoma Hazemba

**Author notes:** **Corresponding author:** (II).

## Abstract

**Background:** Unintended adolescent pregnancies remain a public health challenge in Zambia, where LARC use is low despite their effectiveness. Barriers such as stigma, misconceptions, and limited access persist. Previous studies conducted in Lusaka district did not explore the challenges faced by adolescents in trying to access and use LARCs. Understanding these challenges is crucial for developing targeted interventions to promote safe and effective contraception practices. This study examined factors influencing adolescent knowledge, willingness, and uptake of LARCs in public health facilities in Lusaka.

**Methods:** A cross-sectional study was conducted between *November 2024 and March 2025* among adolescent girls aged 15–19 years in five first-level hospitals in Lusaka, Zambia, using structured questionnaires to obtain quantitative data^1^. LARC use was measured as a binary outcome, with multiple regression identifying associated factors.

**Results:** There were 400 participants in total, of whom 48.0% (181/377) reported ever using a LARC. In the investigator-led adjusted key-predictor model, age was associated with higher odds of ever using LARC (AOR = 1.31, 95% CI: 1.01-1.69; *p* = 0.042). Adolescents who were not willing to delay pregnancy had higher odds of LARC use than those willing to delay pregnancy (AOR = 4.31, 95% CI: 1.05-17.73; *p* = 0.043). Knowledge of LARCs (AOR = 2.81, 95% CI: 1.27-6.19; *p* = 0.011), having children (AOR = 2.71, 95% CI: 1.32-5.54; *p* = 0.006), and residence in Chipata compared with Kanyama (AOR = 231.96, 95% CI: 28.51-1887.37; *p* < 0.001) were also significant predictors..

**Conclusions:** The findings indicate that older age, knowledge of LARCs, reproductive experience, fertility intentions, and facility context were associated with ever use of LARCs. The large residence effect points to clinic-level service delivery differences, highlighting the need for targeted strategies to improve equitable access to LARCs, particularly among younger women and facilities with lower uptake, while future research should investigate service delivery factors and determinants of current LARC use.

## Introduction

Unintended pregnancies among adolescents remain a major public health concern globally, and more so in sub-Saharan Africa. These pregnancies often result in severe health and social consequences, including maternal mortality due to unsafe abortions, psychological distress such as depression and suicide, and interruptions to education and future economic opportunities (Sambah et al., 2022). Adolescents, defined as individuals aged 10 to 19 years (Te et al., 2025), constitute a significant and rapidly growing segment of the global population (Christian & Smith, 2018; Gore et al., 2011)^4^. In Zambia, teenage pregnancy remains a persistent challenge, with the 2024 Zambia Demographic and Health Survey (ZDHS, 2024) reporting a prevalence of 28.4% in rural areas and 13.2% in urban settings. The urgency of ensuring access to effective reproductive health services, particularly for this vulnerable population group, cannot be over emphasized.

Long-acting reversible contraceptives (LARCs), which include intrauterine devices (IUDs) and implants, offer a highly effective solution to prevent unintended pregnancies, especially among adolescents. These methods are over 99% effective, have low discontinuation rates, and do not rely on consistent user action, making them ideal for young people who may struggle with regular contraceptive use (Ochako et al., 2015; Yinger, 2016). In countries like the United States, increased uptake of LARCs has been associated with dramatic reductions in unintended pregnancies and abortions (Shoupe, 2016). Additionally, LARCs provide non-contraceptive benefits such as alleviating menstrual disorders and improving hemoglobin levels (Yinger, 2016). Despite these advantages, the use of LARCs among adolescents in Zambia remains remarkably low. The 2024 ZDHS (2024) reports a contraceptive prevalence of only 0.2% for IUDs and 8.1% for implants among girls aged 15–19.

In Zambia, overall contraceptive use has steadily increased among reproductive-aged women over the past three decades; however, adolescents consistently remain the group with the lowest uptake. (Phiri et al., 2024) showed that contraceptive prevalence among sexually active women rose from 14.2% in 1992 to 45.0% in 2018, with behavioural change, rather than demographic shifts, accounting for up to 85% of the increase. These changes were strongly associated with educational attainment, reduced fertility intentions, and exposure to family planning messages.

Similarly, (Chola et al., 2023) found that changes in contraceptive use among adolescent girls were driven more by behavioural patterns than compositional characteristics, highlighting persistent disparities linked to age, education, marital status and rural residence. Their decomposition analysis demonstrated that adolescents aged 15–19 continue to have significantly lower modern contraceptive use compared to older women, despite high awareness. These findings suggest that adolescent contraceptive behaviour is shaped not only by knowledge and access but also by social context and perceived readiness for pregnancy prevention.

Previous studies on contraceptive use have primarily focused on general populations or on short-acting methods such as injectables and pills (Bakibinga et al., 2019; Ahinkorah, 2020). Although some research has addressed adolescent contraceptive behavior; few studies have examined the specific factors influencing the uptake of LARCs among adolescents in Zambia. Existing data from programs such as Performance Monitoring for Action (MOH, 2022) provide prevalence rates but do not explore the underlying causes of low LARC use. Factors such as provider bias, where healthcare providers limit access based on a client’s age, marital status, or parity, remain poorly understood (Cooper et al., 2017; De Vargas Nunes Coll et al., 2019). In addition, adolescents often harbor misconceptions about LARCs, such as fears about infertility, side effects, or delayed return to fertility, which further deter use (Adedini, Omisakin and Somefun, 2019; Ochako et al., 2015).

Filling this gap is very important. Without a deeper understanding of the social, cultural, and health system factors influencing LARC use among adolescents, policies and interventions risk being misaligned with the actual needs of young people. Addressing these gaps can support the design of adolescent-friendly services, eliminate provider bias, and enhance accurate contraceptive counselling. This would not only reduce the incidence of unintended pregnancies but also contribute to better educational and economic outcomes for adolescent girls, ultimately advancing national development goals (Solo and Festin, 2019).

This study therefore aims to investigate the factors influencing the uptake of long-acting reversible contraceptives among adolescent girls attending Family Planning Clinics located within first-level hospitals in Lusaka, Zambia. In the Zambian health system, first-level hospitals are public health facilities that provide outpatient services, basic emergency care, and reproductive health services, including free access to LARC methods under the national family planning program. These facilities were therefore selected as the primary point of contact for adolescents seeking contraceptive services. By exploring social, cultural, and health-service-related determinants of LARC use, the research seeks to generate evidence that can inform policy and programmatic strategies to improve access, acceptability, and utilization of LARCs among adolescents. The ultimate goal is to enhance reproductive health outcomes, reduce teenage pregnancies, and empower young women to make informed choices about their reproductive lives.

## Methods and Materials

### Study design and sampling

This was a cross-sectional study conducted at Family Planning Clinics based within five first-level hospitals in Lusaka District, Zambia. First-level hospitals are public health facilities that provide outpatient care, basic emergency services, and reproductive health services, including contraceptive counseling and LARC provision. These facilities were purposively selected because they offer family planning services free of charge under Zambia’s national reproductive health policy and represent the primary point of access for adolescents seeking contraceptive care. Therefore, they provide an appropriate setting for examining factors influencing LARC uptake among adolescent girls.

The sample size for the study was calculated using the prevalence of 50%. This prevalence was used in order to ensure that the largest sample size is calculated and reduce the risk of underpowering the study. This calculation was intended primarily to estimate the prevalence of LARC use with adequate precision; the regression analysis was therefore interpreted using effect estimates and 95% confidence intervals rather than as a sample size calculation designed around one prespecified odds ratio.

The following formula was used:

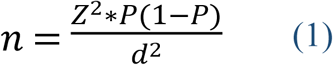

*Where: n is the minimum sample size, Z is the standard normal deviation, set at 1.96, p is the prevalence of modern contraceptive use at 50% (0.5), and q is 1−p (0.5). The degree of accuracy was set at 0.05*.

The sample size was calculated to be 384 adolescents between the ages of 15 - 19, with a 10% attrition rate. The study employed a cluster sampling method to ensure representation from all five hospitals. Each hospital was considered to be a cluster, and within each cluster, adolescents were randomly selected for participation using a sampling frame and a sampling interval of 4. This allowed for efficient data collection while maintaining diversity in responses. Although cluster sampling by facility was used operationally to obtain participants from all five hospitals, the achieved sample size was not inflated by a design effect. Facility/residence was therefore retained in the adjusted analysis and this limitation is acknowledged.

Despite the calculated sample size being 384, more women were willing to be interviewed. Hence the final study involved 400 women who presented at the first level hospitals in Lusaka, Zambia.

### Data collection

Data were collected using a structured questionnaire, between 1^st^ November 2024 and 30^th^ March 2025. This questionnaire was developed based on existing literature and expert input. The questionnaire was pre-tested at one of the first-level hospitals that was not included in the final study to ensure clarity and reliability. It covered various themes, including demographic characteristics, knowledge, and perceptions about LARCs, accessibility, and barriers to use. Trained research assistants administered the questionnaires in a face-to-face setting to ensure accuracy and completeness of responses. The full questionnaire used for data collection is provided in the Appendix for reference.

Adolescents between the ages 15-19 years old (inclusive) at the time of the study, able to understand and respond to questions about sexual and reproductive health in English or the local language spoken fluently and were able to provide informed consent or assent if below 19 years old were recruited in one of the examination rooms at each facility. The study excluded all adolescents with cognitive impairments that would hinder their ability to provide informed consent or participate actively in the study.

Questionnaires were administered to adolescents who visited the Family Planning Clinics by trained nurses of the clinic. During data collection, adolescents were interviewed individually in a private setting. Parents or guardians were not present during questionnaire administration to ensure confidentiality and reduce response bias; however, informed consent and assent procedures were strictly followed in accordance with ethical guidelines. Because nurses working in the study clinics administered the questionnaire, social desirability bias may have occurred, especially for questions on contraceptive use, knowledge, and willingness to use LARCs.

The questionnaire was divided into two sections: The first section obtained information on sociodemographic characteristics such as age, education, number of living children, and utilization of modern contraception. The sexual health history section gathered information on participants’ age at first sexual encounter, pregnancy and birth history, HIV status and treatment, willingness to delay pregnancy, and past contraceptive use, including the types of contraceptives previously used.

The second section assessed knowledge, attitude and factors affecting the use of LARC methods. The questions assessing knowledge of specific LARC types (implants and intrauterine devices); injectables were treated as non-LARC contraceptive methods, understanding of their use, and perceptions of their effectiveness in preventing pregnancy. Attitudes towards LARCs explored participants’ willingness to consider LARCs as a contraceptive method, reasons for their preferences (including perceived convenience or concerns such as discomfort and fear of pain), and their willingness to pay for LARC services if offered. While experiences with LARCs were designed to capture prior use, the type of LARC used sources of information about LARCs, and any challenges encountered in accessing or initiating their use, such as cost, availability, or lack of service providers. Utilization of long-acting contraceptive methods among the women was set as binary outcome variable. The questionnaire did not include a full method information index, and this is now acknowledged as a limitation.

### Definition of variables

#### The outcome variable

The dependent variable was ever use of a LARC, defined as self-reported past or current use of an implant or intrauterine device. It was coded 1 for adolescents who had ever used an implant or IUD and 0 for adolescents who had never used a LARC. Current use of any contraceptive method was treated separately from ever LARC use.

#### Independent variables

These included challenges to accessing contraceptives were categorized into five groups: (1) Expensive, (2) Not readily available, (3) Lack of service providers, (4) Other, and (5) N/A. Accessibility was classified as (1) Yes and (2) No, indicating whether adolescents reported having access to contraceptive services. Information sources were grouped into six categories: (1) Healthcare provider, (2) Friends, (3) School/community programs, (4) Internet/social media, (5) Other, and (6) More than one source.

The type of contraceptive method used was coded into four categories: (1) Injectable, (2) Implant, (3) Intrauterine device (IUD), and (4) N/A for those who did not specify or were not using any method. Current use of any contraceptive method was categorized separately from the dependent variable for ever LARC use. Payment for contraceptive services was classified as (1) Yes and (2) No, reflecting whether individuals had to pay for their chosen method.

Reasons for not using contraceptives were categorized into four groups: (1) Risk of infection, (2) Uncomfortable, (3) Fear of pain, and (4) N/A. The primary reason for choosing a contraceptive method was categorized as (1) Convenient, (2) Easy to use, (3) Other, (4) N/A, and (5) More than one reason. Willingness to use Long-Acting Reversible Contraceptives (LARC) was coded into three categories: (1) Yes, (2) No, and (3) Not sure. Perceived effectiveness of contraceptive methods was categorized into (1) Very effective, (2) Somewhat effective, (3) Not effective, (4) Unsure, and (5) More than one.

Contraceptive method patterns included injectables, implants, IUDs, and more than one method; however, LARC-specific analyses were restricted to implants and IUDs. Awareness of LARC was retained in its original categories: (1) Yes and (2) No. Knowledge of LARC was similarly categorized as (1) Yes and (2) No. The type of contraceptive known was classified into (1) Pills, (2) Condoms, (3) Implants, (4) Loop, (5) Injectable, (6) Others, (7) N/A, and (8) More than one.

Contraceptive use was categorized into (1) Yes and (2) No, indicating whether an individual was using any method at the time of the survey. Delay in seeking contraceptive services was also classified as (1) Yes and (2) No. HIV status was coded into two categories: (1) Positive and (2) Negative, reflecting self-reported or tested status.

#### Data analysis

Data were analyzed using STATA version 17. Prior to analysis, the dataset was assessed for missing values, and records with incomplete responses on key variables were excluded using complete-case (listwise) analysis to ensure data integrity. The first step involved extracting a dataset of adolescents from 400 questionnaires for participants aged 15–19 years. Descriptive statistics were then used to profile the study population based on selected sociodemographic and reproductive health characteristics.

Next, bivariate analysis was conducted to examine associations between independent variables and LARC use, using Pearson’s Chi-squared test (χ²) to determine statistical significance at a 95% confidence level (*p* < 0.05). Cross-tabulation was also used to explore distribution patterns across categories.

To determine factors associated with LARC uptake, binary logistic regression was applied because the dependent variable, ever used LARC, was coded as a binary variable (Yes/No). Crude associations were presented as univariate results and were not interpreted as final evidence. The primary investigator-led adjusted model included age, willingness to delay pregnancy, LARC knowledge, having children, and residence/facility context. These variables were selected to reflect key predictors while avoiding over-adjustment for variables likely to lie on the reproductive pathway. A *p*-value of <0.05 was considered statistically significant.

#### Ethical considerations

This study obtained ethical approval from The University of Zambia Biomedical Research and Ethics Committee (UNZABREC) with approval number: REF.NO.5449-2024. Further approval was obtained from the National Health and Research Authority (NHRA) with approval number : NHRA-1372/14/07/2024. Written informed consent was obtained from all participants before data collection. Confidentiality and anonymity were strictly maintained throughout the study. Participants were informed of their right to withdraw from the study at any time without any consequences.

## Results

### Characteristics of the study population

The first stage of the analysis profiled the study group against the selected variable characteristics, and the results are presented in table 1.

**Table 1.** Background characteristics of adolescents aged 15 – 19 years old.

| Characteristics | Number | Percentage (%) |
| --- | --- | --- |
| **Age** |  |  |
| 15 | 15 | 3.8% |
| 16 | 35 | 8.8% |
| 17 | 76 | 19.0% |
| 18 | 126 | 31.6% |
| 19 | 147 | 36.8% |
| Total | 399 | 100.0% |
| **Religious Affiliation** |  |  |
| Catholic | 104 | 26.2% |
| Pentecostal | 138 | 34.8% |
| Protestant | 75 | 18.9% |
| Other | 80 | 20.2% |
| Total | 397 | 100.0% |
| **Education** |  |  |
| Primary | 125 | 31.3% |
| Secondary | 223 | 55.9% |
| Tertiary | 29 | 7.3% |
| None | 22 | 5.5% |
| Total | 399 | 100.0% |
| **Marital Status** |  |  |
| Never married | 215 | 54.3% |
| Cohabiting/ Married | 171 | 43.2% |
| Formerly married | 10 | 2.5% |
| Total | 396 | 100.0% |
| **Age at first sexual encounter** |  |  |
| 12 | 4 | 1.0% |
| 13 | 9 | 2.3% |
| 14 | 34 | 8.6% |
| 15 | 109 | 27.5% |
| 16 | 122 | 30.8% |
| 17 | 76 | 19.2% |
| 18 | 28 | 7.1% |
| 19 | 14 | 3.5% |
| Total | 396 | 100.0% |
| **Number of pregnancies** |  |  |
| 1 | 132 | 33.4% |
| 1 | 174 | 44.1% |
| 2 | 70 | 17.7% |
| 3 | 16 | 4.1% |
| 4 | 2 | 0.5% |
| 9 | 1 | 0.3% |
| Total | 395 | 100.0% |
| **HIV Status** |  |  |
| Positive | 65 | 16.5% |
| Negative | 281 | 71.5% |
| Unknown | 47 | 12.0% |
| Total | 393 | 100.0% |
| **Delay pregnancy** |  |  |
| Willing | 373 | 96.9% |
| Not willing | 12 | 3.1% |
| Total | 385 | 100.0% |
| **Ever Used LARC** |  |  |
| Yes | 181 | 48.0% |
| No | 196 | 52.0% |
| Total | 377 | 100.0% |
| **Perceptions about LARC effectiveness** |  |  |
| Effective | 224 | 57.7% |
| Somewhat effective | 50 | 12.9% |
| Not effective | 14 | 3.6% |
| Unsure | 100 | 25.8% |
| Total | 388 | 100.0% |
| **Teenagers aware of LARC** |  |  |
| Yes | 327 | 83.2% |
| No | 66 | 16.8% |
| Total | 393 | 100.0% |
| **Teenage willingness to Use LARC** |  |  |
| Yes | 310 | 78.5% |
| No | 77 | 19.5% |
| Unsure | 8 | 2.0% |
| Total | 395 | 100.0% |
| **Ever used LARC (Uptake)** |  |  |
| Yes | 181 | 48.0% |
| No | 196 | 52.0% |
| Total | 377 | 100.0% |
| **Type of LARC used** |  |  |
| Implant | 108 | 65.9% |
| IUD | 4 | 2.4% |
| 2 or more types | 52 | 31.7% |
| Total | 162 | 100.0% |
| **Residence** |  |  |
| Kanyama | 82 | 20.6% |
| Chawama | 79 | 19.8% |
| Chilenje | 80 | 20.1% |
| Chipata | 82 | 20.6% |
| Matero | 76 | 19.0% |
| Total | 399 | 100.0% |

The study included adolescents aged 15–19 years. The largest proportion were aged 19 years (36.84%), followed by those aged 18 years (31.58%), whereas 15-year-olds made up the smallest proportion (3.76%). The distribution across residential compounds was relatively even: Kanyama and Chipata each accounted for 20.55% of respondents, followed by Chilenje (20.05%), Chawama (19.80%) and Matero (19.05%).

Regarding religion, Pentecostals comprised the largest group (34.76%), followed by Catholics (26.20%), other religions (20.15%) and Protestants (18.89%). Most adolescents had secondary education (55.89%), while 31.33% had only primary education, 7.27% had tertiary education and 5.51% had no formal education. More than half of participants were never married (54.29%), with 43.18% married or cohabiting and 2.53% divorced, separated or widowed.

Most adolescents reported sexual debut between ages 15 and 17. HIV status was predominantly negative (71.50%), while 16.54% were HIV-positive. Nearly all respondents (96.88%) indicated willingness to delay pregnancy. Awareness/knowledge of LARCs was high, and 78.48% expressed willingness to use them. Overall, 48.0% reported ever using a LARC. After removing injectables from the LARC-specific method category, implants were the most frequently reported LARC method, while IUD use was uncommon. One participant reported nine pregnancies before age 19; this uncommon self-reported value was checked in the dataset and retained, but interpreted cautiously.

### Differentials in LARC Use

Long-Acting Reversible Contraceptives (LARCs) offer an effective and convenient form of birth control, yet their uptake among adolescents remains low. This study examines the socio-demographic factors, knowledge, attitudes, and challenges influencing LARC use among adolescents in Lusaka, Zambia. The findings are based on cross-tabulations and statistical associations derived from rank-sum and chi-square tests shown in Table 2 below.

**Table 2.**
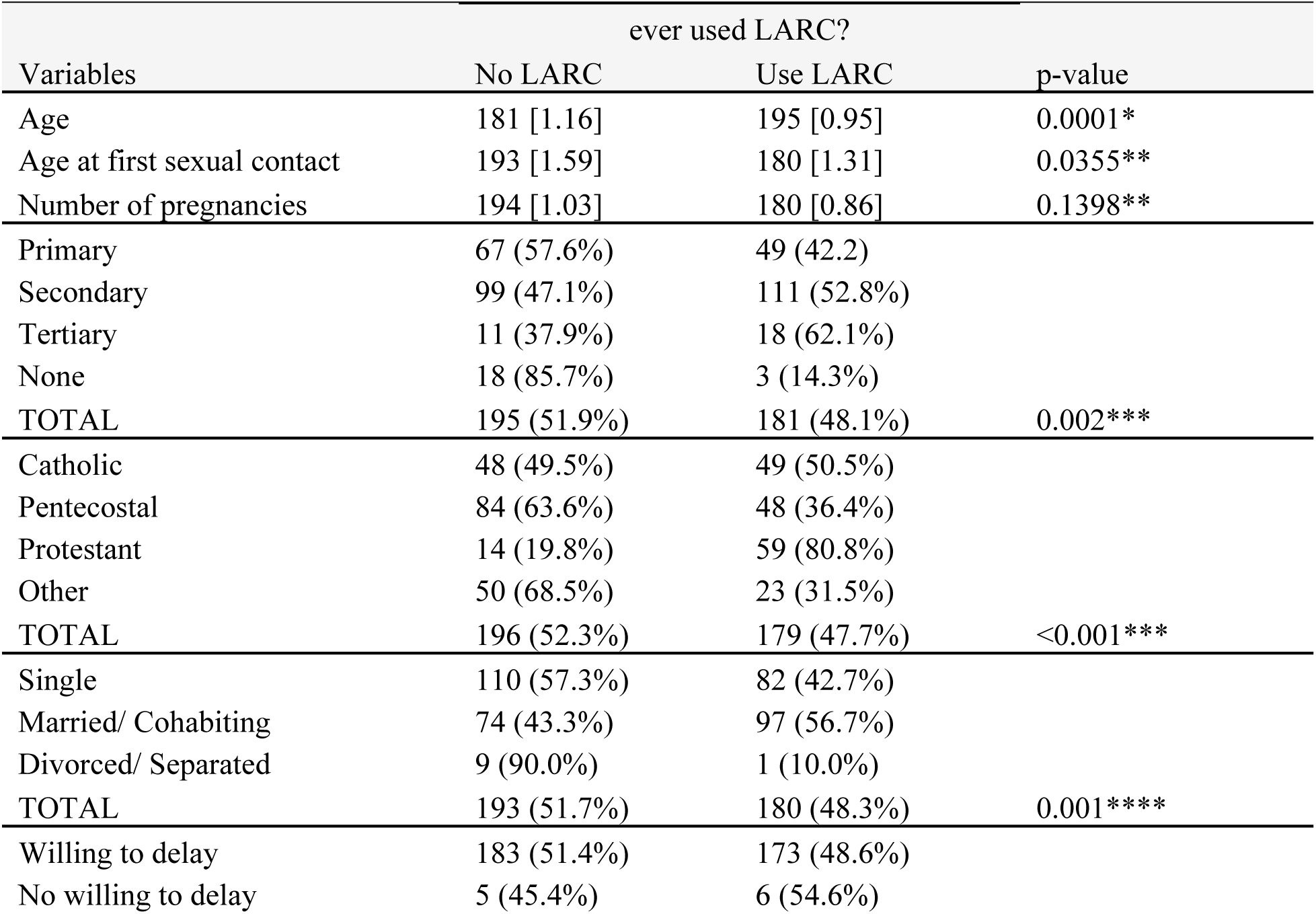
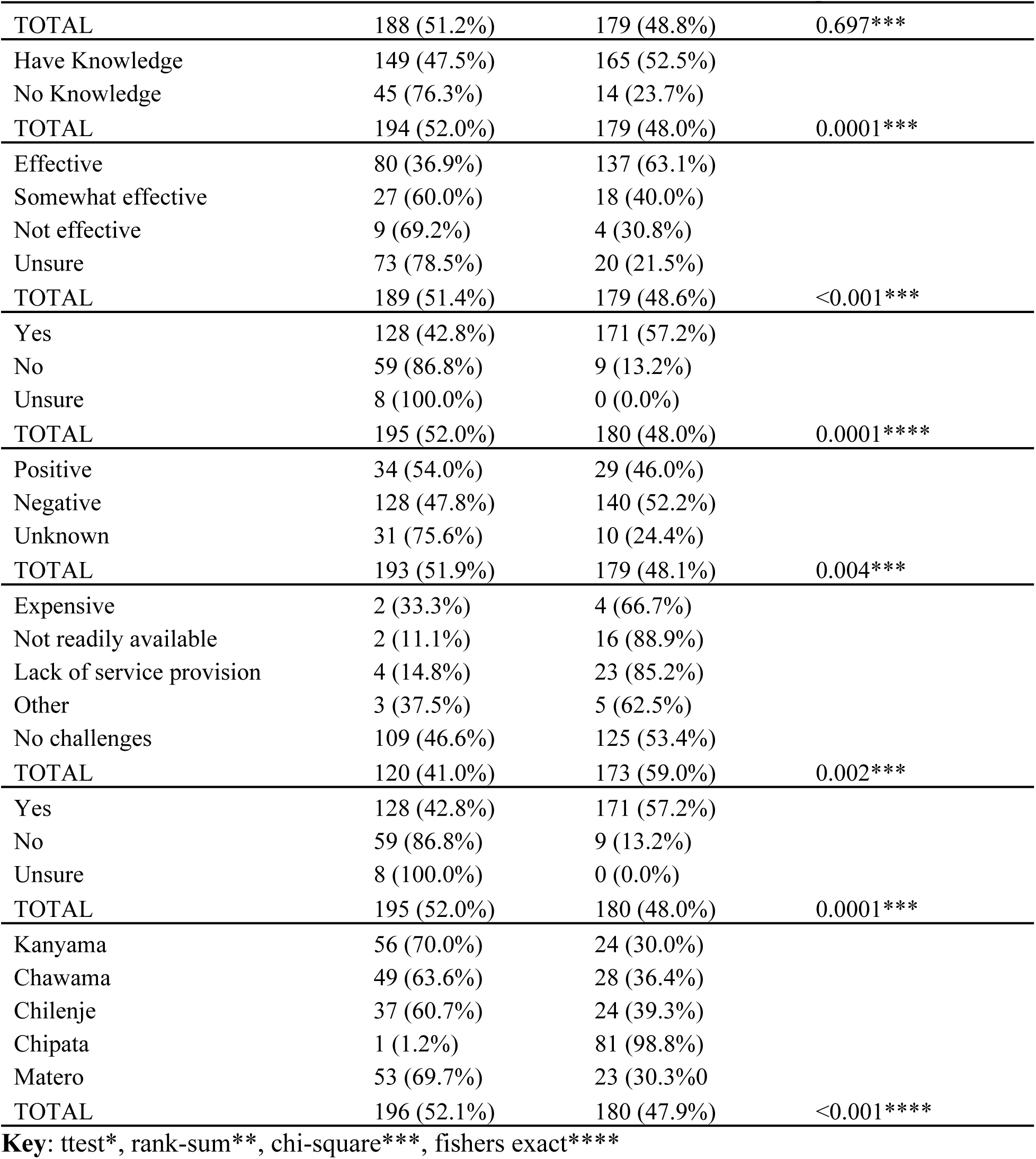
Differentials of LARC use for adolescent girls aged 15-19 years: Measures of associations.

Adolescents with secondary or tertiary education were significantly more likely to use LARCs than those with little or no education (*p*=0.002). Religion showed a strong association with LARC use (*p*=0.0001), with Protestants having the highest uptake and Pentecostals the lowest. Residence was also significant (*p*=0.000), with adolescents in Chipata reporting the highest usage (98.78%) compared with lower rates in Kanyama, Chawama, Chilenje, and Matero. Marital status influenced uptake (*p*=0.001), as married or cohabiting adolescents were more likely to use LARCs than those who were single or separated. HIV status was also significant (*p*=0.004); while HIV-negative adolescents had higher usage, those with unknown status demonstrated the highest willingness to use LARCs. Willingness to delay pregnancy did not show a significant association with LARC use (*p*=0.697).

Adolescents who had knowledge of LARCs were more likely to have used them (52.55%) than those without knowledge (23.73%) (*p*=0.0001). Positive perceptions of LARC effectiveness were associated with greater uptake (63.13% versus 30.77% among those who perceived them as ineffective) (*p*=0.0001). Willingness to use LARCs showed a strong association with actual use, as 57.19% of willing adolescents had used a LARC compared to only 13.4% of those who rejected them (*p*=0.0001), while those who were unsure or unwilling showed minimal or no uptake.

### Factors Influencing Ever Use of Long-Acting Reversible Contraception

This analysis examined the socio-demographic and reproductive health factors associated with ever use of long-acting reversible contraceptives (LARCs) among adolescents. Both univariate and multivariable logistic regression analyses were conducted to assess the relationship between each explanatory variable and the outcome of interest. Crude and adjusted odds ratios (ORs), 95% confidence intervals (CIs), and p-values were used to determine the strength and statistical significance of these associations. The findings are presented in Table 3.

**Table 3.** Factors associated with use LARC (LARC uptake)

| Variables | Crude Regression |  | Adjusted Regression |  |
| --- | --- | --- | --- | --- |
|  | OR (95% CI) | p-value | AOR (95% CI) | p-value |
| Age | 1.623 (1.32 - 1.99) | <0.001 | 1.31 (1.01 - 1.69) | 0.042 |
| **Education** |  |  |  |  |
| No education | 1.00 |  |  |  |
| Primary | 4.39 (1.22 - 15.73) | 0.023 |  |  |
| Secondary | 6.73 (1.92 - 23.53) | 0.003 |  |  |
| Tertiary | 9.82 (2.34 - 41.19) | 0.002 |  |  |
| **Religion** |  |  |  |  |
| Catholic | 1.00 |  |  |  |
| Pentecostal | 0.55 (0.32 - 0.94) | 0.028 |  |  |
| Protestant | 4.13 (2.04 - 8.36) | <0.001 |  |  |
| Other | 0.46 (0.24 - 0.85) | 0.014 |  |  |
| **Marital status** |  |  |  |  |
| Single | 1.00 |  |  |  |
| Married | 1.76 (1.16 - 2.67) | 0.008 |  |  |
| Formerly married | 0.15 (0.18 - 1.20) | 0.074 |  |  |
| Age 1st sexual contact | 0.89 (0.77 - 1.03) | 0.115 |  |  |
|  | OR (95% CI) | p-value | AOR (95% CI) | p-value |
| Number of pregnancies | 0.84 (0.67 - 1.05) | 0.129 |  |  |
| **HIV status** |  |  |  |  |
| Positive | 1.00 |  |  |  |
| Negative | 1.28 (0.74 - 2.22) | 0.376 |  |  |
| Unknown | 0.38 (0.16 - 0.90) | 0.028 |  |  |
| **Delay pregnancy** |  |  |  |  |
| Yes | 1.00 |  | 1.00 |  |
| No | 1.27 (0.38 - 4.24) | 0.698 | 4.31 (1.05 - 17.73) | 0.017 |
| **Knowledge** |  |  |  |  |
| No | 1.00 |  | 1.00 |  |
| Yes | 3.66 (1.94 - 6.93) | <0.001 | 2.81 (1.27 - 6.19) | 0.011 |
| **Children** |  |  |  |  |
| No children | 1.00 |  | 1.00 |  |
| Has children | 1.03 (0.55 - 1.29) | 0.437 | 2.71 (1.32 - 5.54) | 0.006 |
| **Residence** |  |  |  |  |
| Kanyama | 1.00 |  | 1.00 |  |
| Chawama | 1.33 (0.68 - 2.60) | 0.398 | 1.00 (0.48 - 2.03) | 0.977 |
| Chilenje | 1.51 (0.75 - 3.05) | 0.247 | 2.21 (0.99 - 4.93) | 0.053 |
| Chipata | 189 (24.84 - 1437.87) | <0.001 | 231.96 (28.51 - 1887.37) | <0.001 |
| Matero | 1.01 (0.51 - 2.01) | 0.971 | 0.64 (0.31 - 1.34) | 0.236 |

In the crude analysis, age, education, religion, marital status, knowledge of LARCs, HIV status, and residence were significantly associated with ever use of LARCs. The odds of ever using a LARC increased with age (OR = 1.62, 95% CI: 1.32–1.99, *p* < 0.001). Compared with women with no education, those with primary (OR = 4.39, 95% CI: 1.22–15.73, *p* = 0.023), secondary (OR = 6.73, 95% CI: 1.92–23.53, *p* = 0.003), and tertiary education (OR = 9.82, 95% CI: 2.34–41.19, *p* = 0.002) had significantly higher odds of ever using a LARC. Compared with Catholics, Pentecostals had lower odds of LARC use (OR = 0.55, 95% CI: 0.32–0.94, *p* = 0.028), while Protestants had higher odds (OR = 4.13, 95% CI: 2.04–8.36, *p* < 0.001). Married women had higher odds of LARC use than single women (OR = 1.76, 95% CI: 1.16–2.67, *p* = 0.008). Women with knowledge of LARCs were more likely to have ever used a LARC (OR = 3.66, 95% CI: 1.94– 6.93, *p* < 0.001). Residence in Chipata was strongly associated with LARC use compared with Kanyama (OR = 189.00, 95% CI: 24.84–1437.87, *p* < 0.001).

After adjustment, age remained significantly associated with ever LARC use (AOR = 1.31, 95% CI: 1.01–1.69, *p* = 0.042). Women who did not intend to delay pregnancy had higher odds of ever using a LARC than those who intended to delay pregnancy (AOR = 4.31, 95% CI: 1.05–17.73*, p* = 0.017). Knowledge of LARCs remained an independent predictor, with knowledgeable women having nearly three times higher odds of ever use (AOR = 2.81, 95% CI: 1.27–6.19, *p* = 0.011). Women with children were more likely to have ever used a LARC than women without children (AOR = 2.71, 95% CI: 1.32–5.54, *p* = 0.006). Residence remained an important determinant, with women attending the Chipata facility having substantially higher odds of ever LARC use than those attending Kanyama (AOR = 231.96, 95% CI: 28.51–1887.37, *p* < 0.001). No significant associations were observed for Chawama, Chilenje, or Matero after adjustment.

## Discussion

Our study examined factors associated with ever use of long-acting reversible contraception (LARC) among adolescents attending Family Planning Clinics, with 48.0% reporting ever use of a LARC. The investigator-led adjusted model identified age, LARC knowledge, having children, fertility intentions, and residence context as important predictors. These findings suggest that both individual reproductive experience and service-delivery context shape adolescent LARC uptake. The observed prevalence indicates that LARC use is relatively common among adolescents accessing family planning services, although substantial disparities remain across subgroups. Furthermore, the persistence of facility-level differences after adjustment highlights the potential influence of service availability, provider practices, and local program implementation on contraceptive choices.

Socio-demographic factors significantly influenced adolescents’ willingness to use LARCs in Lusaka. Increasing age showed higher use, consistent with findings from Kenya and Uganda linking age to increased contraceptive uptake due to greater reproductive health exposure (Ochako et al., 2015; Kungu, Agwanda and Khasakhala, 2023). Although education was associated with crude odds of uptake, it did not remain significant after adjustment, suggesting that service-related rather than educational factors may influence use in this setting. Similar observations were reported in Ethiopian clinics where education alone did not drive uptake without provider reinforcement (Pazol et al., 2018).

Adolescents unwilling to delay pregnancy are significantly more likely to use LARC, suggesting that reproductive intentions strongly influence contraceptive choices in first-level hospitals. LARC uptake is higher among adolescents who have experienced pregnancy or are ready for childbearing, while those wishing to avoid pregnancy may face barriers such as misconceptions, limited access, or lack of counseling, resulting in lower LARC use overall (Sambah et al., 2022; Zafiro & Rodriguez, 2017). Factors like marital status, number of children, and exposure to postpartum or hospital-based counseling also affect LARC adoption, with those not seeking to delay pregnancy showing greater acceptance(Sambah et al., 2022). Tailored counseling and youth-friendly services are needed to support informed LARC decisions among adolescents with diverse reproductive goals (Shimels et al., 2025).

Despite relatively high awareness, the barriers reported (e.g., cost, availability, and inadequate provision) should be interpreted cautiously, as only a small number of respondents answered this question (n=6 for cost). Nevertheless, they align with evidence from sub-Saharan Africa, where restrictive social norms, provider biases, and misinformation hinder use (Bain, Amu and Tarkang, 2021). Evidence from Malawi and Tanzania also indicates that consistent supply and trained personnel are critical for adoption (Chandra-Mouli et al., 2014), aligning with our findings that residence significantly affected contraceptive uptake. Adolescents from Chipata had substantially higher uptake, mirroring reports of geographic variability in service readiness across Zambia and Kenya (Kamanga et al., 2023; Stonehill et al., 2020).

Importantly, the gap observed between willingness to use LARCs (78.48%) and actual uptake (48%) suggests unrealized demand, a finding similar to studies where adolescents expressed intention but faced structural or service-level constraints (Solo and Festin, 2019a; Duvall et al., 2014). This indicates a missed opportunity to engage adolescents already attending FP clinics, especially as high willingness may serve as a foundation for expanding method provision and counseling. Strengthening adolescent-centered counseling, integrating HIV screening, and ensuring reliable LARC availability may support greater uptake and continuity of care (Kamanga et al., 2023; Shimels et al., 2025).

## Policy and Programmatic Implications

The findings underscore the need for adolescent-friendly reproductive health policies. Integrating LARC counseling into school health programs could enhance knowledge and acceptance. Additionally, expanding community distribution models using peer educators and community health workers could improve accessibility. Programs in Kenya and Nigeria employing this approach have reported increased LARC use among adolescents (Jacobstein, 2018).

Provider training should also be prioritized to address biases and ensure adolescents receive nonjudgmental, evidence-based contraceptive counseling. Evidence from Malawi and Tanzania suggests that when providers receive adolescent-friendly service training, contraceptive uptake increases (Chandra-Mouli *et al*., 2014).

## Study limitations

While this study provides valuable insights into factors influencing LARC use among adolescents in Lusaka, some limitations should be acknowledged. Self-reported data may be subject to recall or social desirability bias. The study included only adolescents already attending Family Planning Clinics who may be more informed or motivated regarding contraception; therefore, the results may not represent adolescents who do not access such services. In addition, the study was limited to first level hospitals in Lusaka and may not be generalizable to rural areas or private health facilities. Despite these limitations, the study offers critical information for policymakers and healthcare providers aiming to improve adolescent contraceptive access and uptake.

Additionally, questionnaire administration by clinic nurses may have increased social desirability bias. The study also did not measure the method information index, limiting assessment of counseling quality and service-related factors. In addition, the sample size was calculated for prevalence estimation and was not inflated for a cluster design effect, so adjusted findings should be interpreted with attention to confidence intervals and facility-level variability.

## Conclusion

Our study highlights the relationship between age, LARC knowledge, reproductive experience, fertility intentions, and facility/residence context in influencing ever use of LARCs among adolescents in Lusaka, Zambia. The revised adjusted model indicates that age, knowledge, having children, not being willing to delay pregnancy, and Chipata residence were associated with LARC uptake, while univariate associations should be interpreted as descriptive rather than final adjusted findings. These findings underscore the need for adolescent-responsive family planning programmes that strengthen LARC counselling, improve contraceptive knowledge, and ensure equitable access across facilities. Future research should investigate facility-level factors and service delivery practices that may explain the substantial differences in LARC uptake observed between clinic settings.

## Data Availability

The minimal data set is available at Dryad via (DOI): https://doi.org/10.5061/dryad.bnzs7h4sp

## Acknowledgements

Pre-Publication Support Service (PREPPS) supported the development of this manuscript.

